# Content analysis and characterization of medical tweets during the early Covid-19 pandemic

**DOI:** 10.1101/2020.12.22.20248712

**Authors:** Ross Prager, Michael Pratte, Rudy R. Unni, Sudarshan Bala, Nicholas Ng Fat Hing, Kay Wu, Trevor A. McGrath, Adam Thomas, Laura Hilary Thompson, Julia Hajjar, Brent Thoma, Philippe Rola, Alan Karovitch, Matthew DF McInnes, Kwadwo Kyeremanteng

## Abstract

**Objective:** The novel coronavirus disease 2019 (Covid-19) has infected millions worldwide and impacted the lives of many folds more. Many clinicians share new Covid-19 related resources, research, and ideas within the online Free Open Access to Medical Education (FOAM) community of practice. This study provides a detailed content and contributor analysis of Covid-19 related tweets among the FOAM community.

**Design, Setting, Participants:** Twitter was searched from November 1^st^, 2019 to March 21^st^, 2020 for English tweets discussing Covid-19 in the FOAM community. Tweets were classified into one of 13 pre-specified content categories: original research, editorials, FOAM resource, public health, podcast or video, learned experience, refuting false information, policy discussion, emotional impact, blatantly false information, other Covid-19, and non-Covid-19. Further analysis of linked original research and FOAM resources was performed. 1000 randomly selected contributor profiles and those deemed to have contributed false information were analyzed.

**Results:** The search yielded 8541 original tweets from 4104 contributors. The number of tweets in each content category were: 1557 other Covid-19 (18·2%), 1190 emotional impact (13·9%), 1122 FOAM resources (13·1%), 1111 policy discussion (13·0%), 928 advice (10·9%), 873 learned experience (10·2%), 424 non-Covid-19 (5·0%), 410 podcast or video (4·8%), 304 editorials (3·6%), 275 original research (3·2%), 245 public health (2·9%), 83 refuting false information (1·0%), and 19 blatantly false (0·2%).

**Conclusions:** Early in the Covid-19 pandemic, the FOAM community used Twitter to share Covid-19 learned experiences, online resources, crowd-sourced advice, research, and to discuss the emotional impact of Covid-19. Twitter also provided a forum for post-publication peer review of new research. Sharing blatantly false information within this community was infrequent. This study highlights several potential benefits from engaging with the FOAM community on Twitter.

## INTRODUCTION

Millions of cases of coronavirus disease 2019 (Covid-19) have been reported globally since the first known case in December 2019.[1-3] Covid-19’s worldwide impact has been recognized through its classification as a global pandemic by the World Health Organization (WHO).[4] Covid-19’s rapid spread has spurred healthcare workers, researchers, and members of the public to search for accurate and up-to-date information online. The rate of new Covid-19-related research, however, has challenged conventional methods of scientific knowledge dissemination (e.g. peer reviewed journals) which do not always publish on rapid timelines.[5] In response, clinicians worldwide have turned to social media to debate new research while sharing their experiences and resources.[6]

Social media use among clinicians is not a new phenomenon. In the past decade an online community of practice has developed with the goal of sharing ideas, research, and learned experiences through freely published online resources.[7,8] Termed “Free Open Access to Medical Education” (FOAM), this movement has become a valuable resource for healthcare professionals and medical learners.[8-11] In addition to relaying explicit medical knowledge it may also be an effective medium for transmitting tacit knowledge (experiential or process based knowledge).[12,13] Compared to traditional peer reviewed journals, FOAM has variable publication and editorial processes relying heavily on post-publication peer review.[14]

Optimizing knowledge translation is important during a pandemic as critical decisions need to be made with limited evidence and potentially practice-changing research can be published at any time. Within the FOAM community, Twitter is the most widely used social media platform to share ideas and discuss new research on Covid-19.[6] On Twitter, contributors generate ‘tweets’ of up to 280 characters in length that can be tagged with searchable hashtags (#) and can include images, website links, and documents. While the important role of Twitter during Covid-19 has been recognized by the scientific community,[6] detailed characterization of its use, strengths, and limitations, including accuracy of content, are needed. The objective of this study was to characterize Covid-19 related Twitter use by the FOAM community, and to describe its content, trends, and contributors. In addition, the potential role of Twitter in spreading misinformation was assessed. This research represents an important first step in evaluating Twitter as a platform for knowledge translation during rapidly evolving healthcare crises.

## METHODS

Research ethics board approval for research involving publicly available data is not required at our institutions. Our protocol was registered on the Open Science Framework (OSF) prior to initiation of data collection (https://osf.io/3tx96/). The original data is also published on OSF. The study has been reported in keeping with the Strengthening the Reporting of Observational Studies in Epidemiology (STROBE) statement.[15] Patients or the public were not involved in the design, conduct, reporting, or dissemination plans of our research.

### Search strategy

We searched Twitter on March 21^st^, 2020 for tweets with relevant hashtags from November 1^st^, 2019 to March 21^st^, 2020 using a commercially available hashtag collating tool, Tweet Binder (Pamplona, Spain). Hashtags were selected by consensus of the authors, several of whom were clinician members of the FOAM community. Tweets were included if they contained both a hashtag commonly used by healthcare professionals to discuss FOAM topics (#FOAMed or #meded or #POCUS or #FOAMcc or #medtwitter), and a hashtag used to discuss Covid-19 (#Covid19 or #coronavirus or #Covid or #Covid-19). Alternatively, two Covid-19 FOAM specific hashtags were also included (#Covid4MDs or #CovidFOAM). The search strategy was not case sensitive.

### Tweet analysis

We extracted the following data: total number of original tweets (original text or image content), retweets (a reposted tweet without modification), reach (number of unique people who saw the tweet), impressions (number of times a tweet was liked or retweeted), total number of contributors (accounts creating tweets), and median original tweets per original contributor.

The content of all original, English-language tweets was analyzed independently by one of 5 authors (MP, SB, KW, NN, RP). To assess interrater reliability, a duplicate extraction of 100 tweets was performed by all extractors. We assigned each tweet to one of 13 pre-determined ‘content categories’ created after consensus discussion between authors: 1) peer reviewed original research study related to Covid-19, excluding editorials, commentaries, or perspective articles; 2) editorial, commentary, or perspective article published in a journal or repository relevant to Covid-19; 3) FOAM resources pertaining to the care of Covid-19 patients; 4) public health agency website or university website (eg. Centers for Disease Control and Prevention); 5) medical podcast or video relevant to Covid-19; 6) personal or learned experience caring for Covid-19 patients; 7) a statement or discussion refuting blatantly false or misleading information regarding Covid-19; 8) a discussion about policy or public health measures related to Covid-19; 9) a discussion of the personal or emotional impact of Covid-19; 10) a tweet that provided blatantly false or misleading information (these were flagged and reviewed in duplicate by two senior authors [RP and RU] to provide consensus on this classification); 11) a tweet asking for advice, or for others to share experience caring for Covid-19 patients; 12) other Covid-19 related tweets that did not fit in the other categories; 13) non-Covid-19 related tweets. The overall best fitting category was selected if multiple classifications were possible, and consensus discussion was allowed if needed. Once categorized, we calculated the number and percentage of tweets in each content category by day and week.

### Contributors, original research, and FOAM content

We determined the demographics of Covid-19 FOAM contributors by reviewing 1000 random profiles of the contributors whose Tweets were captured in the search strategy, extracting: the number of followers, total tweets, country of residence, and contributor source (institution, nurse, staff physician, resident physician, medical student, respiratory therapist, pharmacist, other healthcare professional, non-healthcare professional, non-clinician researcher, healthcare-related group). The profiles were randomized and selected using the randomize function in Microsoft Excel. We performed a similar analysis on the contributor profiles whose tweets were flagged as blatantly false or misleading.

To evaluate the dissemination of original research via Twitter, we analyzed the journal of publication, country of corresponding author, article type (epidemiological study, intervention study, diagnostic study, basic science, case series or other), and the median number of days between online publication (either pre-publication or online) and tweet for each included article. To ensure a focus on new research being conducted on Covid-19 (as opposed to previous coronavirus infections), we excluded research articles published before 2020.

We also identified tweets that linked to FOAM resources and the source (website), type of resource, number of tweets including the resource, and the median number of days between FOAM publication online and the tweet.

### Data analysis

We saved the extracted data in Microsoft Excel 2013 (Washington, USA) and analyzed it using R version 3.6.2 (R Project for Statistical Computing). When appropriate, we assessed the distribution of our data using a Shapiro-Wilks test and calculated the mean (+/- standard deviation) for normally distributed data and median (1^st^ and 3^rd^ interquartile range) for data that was not normally distributed. A post-hoc Mann-Whitney U test was performed to compare the days between publication of original research or FOAM resource and tweet. A post-hoc Mann-Kendall trend test with a Bonferroni correction was used to assess for a trend in percentage of total tweets represented by each content category per week. Interrater reliability was assessed using a Fleiss’ Kappa. Statistical significance was set at a p-value of less than or equal to 0·05.

## RESULTS

The first Tweet matching the search criteria was on January 19^th^, 2020, and from then until to March 21^st^, 2020, 74,758 original tweets and retweets from 52,917 contributors were created. Of these, 8819 (11·8%) were original tweets created by 4104 contributors, and 65,490 (88·2%) were retweets (Table 1). We excluded 278 tweets because they were not written in English or contained broken links. Of the remaining 8541 (11·4%) original tweets, 5494 were stand-alone tweets, 1039 were replies, and 2008 were retweets with comments (Figure 1). The original tweets and retweets reached 95,072,663 Twitter users and had a total of 388,701 likes or replies. Contributors to original tweets had a median number of 489 (IQR 144, 1601) followers and published a median of 1 (IQR 1, 2) original tweets. The Shapiro-Wilks test rejected the null hypothesis of normal distribution for the number of followers per contributor, number of original tweets per original contributor, and number of days between online publication and tweet for FOAM resources or studies p value of < 0·0001.

**Table 1:** Characteristics of total tweets, retweets, and contributors

| Characteristics | Number (%) |
| --- | --- |
| Original Tweets and Retweets | 74 758 (100%) |
| Retweets without Comments | 65 940 (88.2%) |
| Original Tweets | 8 541 (11.4%) |
| Tweets | 5 449 |
| Replies | 1 039 |
| Retweets with comments | 2 008 |
| Median number original tweets per original contributor (IQR) | 1 (1,2) |
| Total Reach (number of unique people who saw the tweet) | 95 072 663 |
| Total Impressions (number of likes and retweets) | 388 701 |
| Total Contributors (tweets and retweets) | 52917 |
| Original Contributors | 4104 |
| Median followers per original contributor (IQR) | 489 (144, 1601) |
| Language (tweets and retweets) |  |
| English | 72 927 (97.6%) |
| Unclassified | 909 (1.2%) |
| Spanish | 560 (0.7%) |
| German | 124 (0.2%) |
| French | 45 (0.1%) |

**Figure 1.**
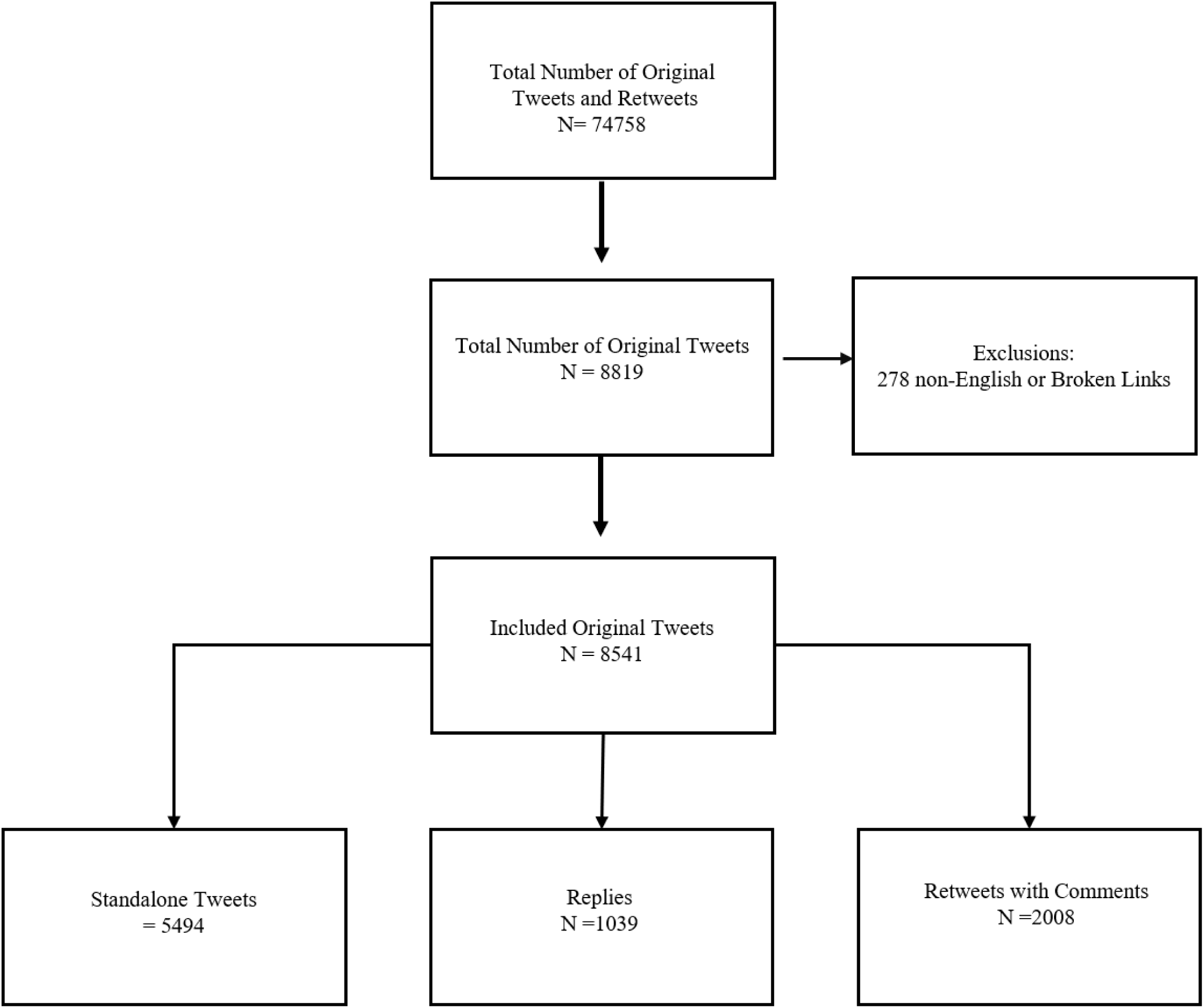
Flow diagram for included tweets.

### Content analysis

The number of tweets per day in each content category are displayed in Figure 2. The tweets per week are displayed in Supplementary Table 1. Most Covid-19-related tweets (1,557 or 18·2%) did not fall into one of the pre-determined categories. There were 1122 tweets sharing FOAM resources including blog posts (13·1%), 275 sharing peer-reviewed original research articles (3·2%), 304 sharing non-research journal articles (editorials, commentaries, or perspectives) (3·6%), 410 sharing podcasts or videos (4·8%), and 245 linking to public health agencies or university websites (2·9%). There were also 1190 tweets discussing emotional impact (13·9%), 1111 tweets discussing public policy (13·0%), 928 tweets asking for advice pertaining to Covid-19 patients (10·9%), and 873 tweets about learned or personal experience caring for Covid-19 patients (10·2%). In addition, there were 83 tweets refuting false information (1·0%), 19 tweets providing blatantly false or misleading information (0·2%) and 424 non-Covid-19 related tweets (5·0%). There was no significant change in the percentage of total tweets represented by each content category over time (see supplementary materials). Overall, interrater reliability was fair with a kappa of 0.31. A selection of representative tweets are provided in Table 2.

**Table 2.**
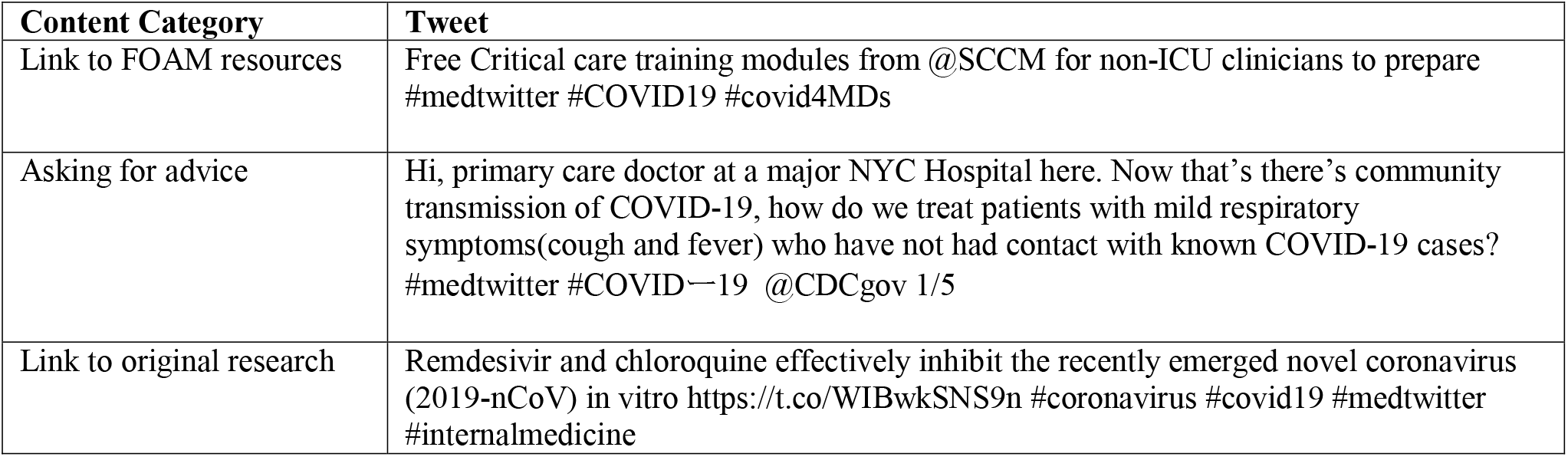
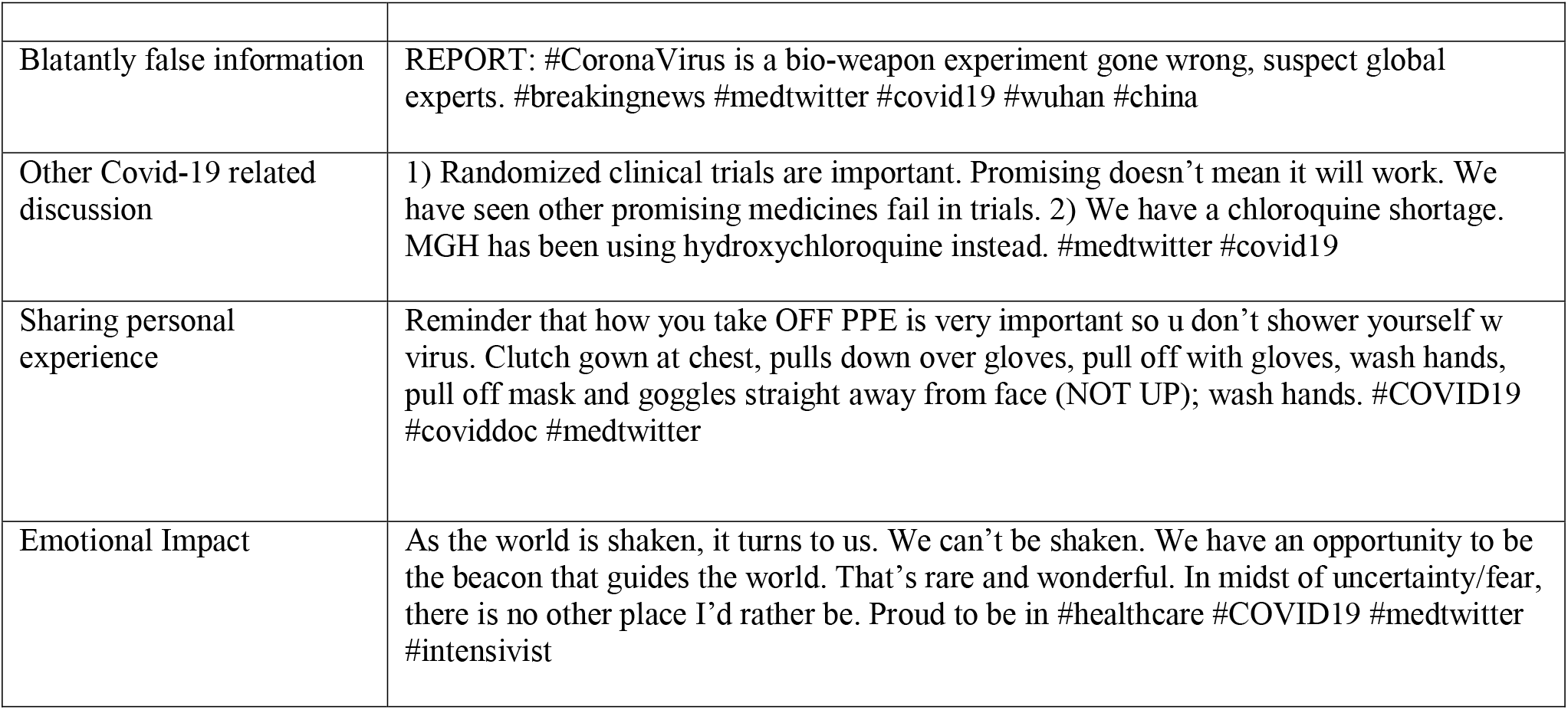
Sample of original tweets included in study

**Figure 2.**
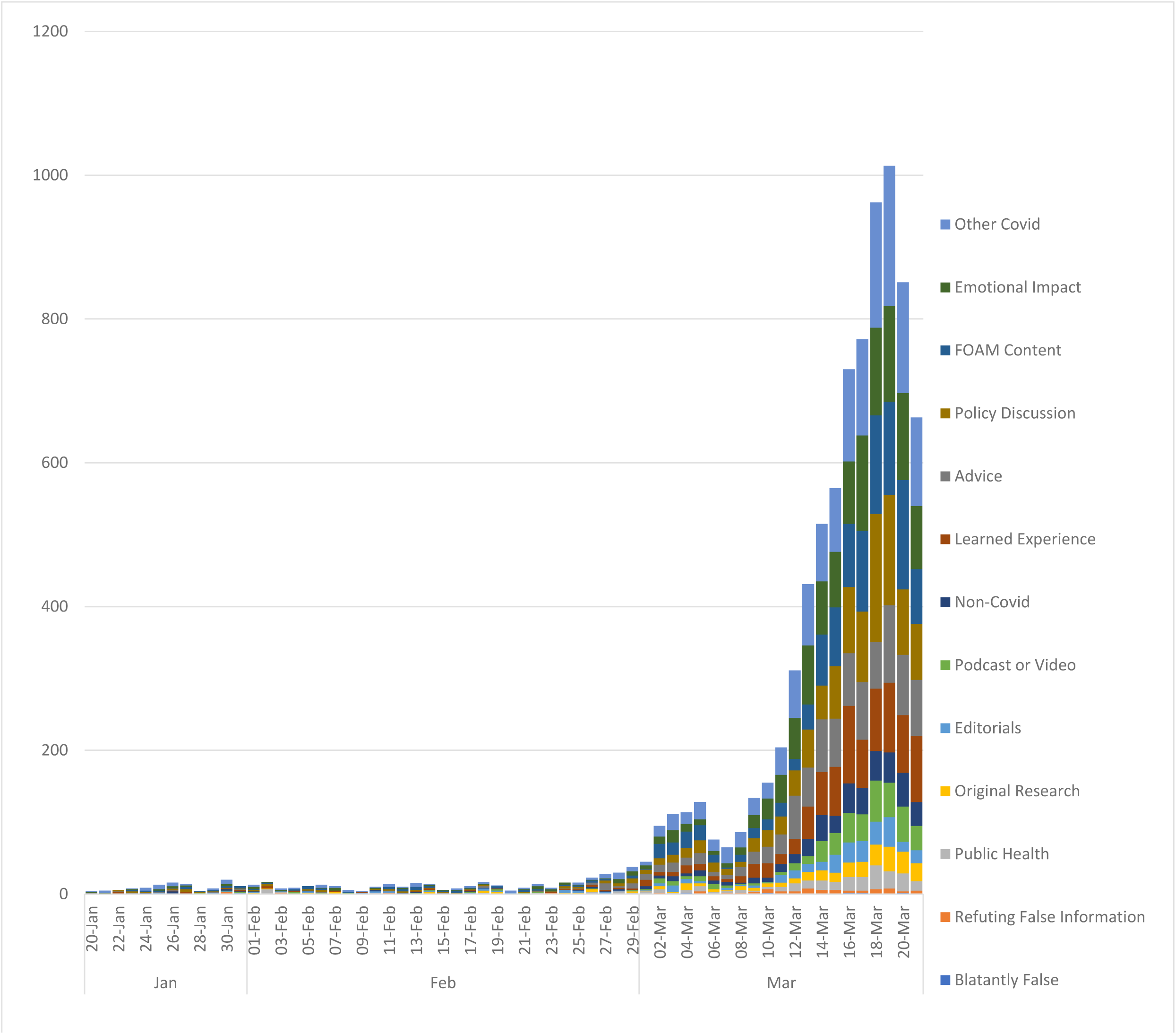
Total number of original tweets by content category per day from January 20^th^ to March 21^st^, 2020.

### Contributor demographics

The 1000 randomly sampled contributors published 2464 tweets which were cast to a total of 2,618,061 followers (Table 3). Their median number of followers was 455 (IQR 136, 1582). Contributors included 437 staff physicians (43·7%), 77 resident physicians (7·7%), 73 non-healthcare professionals (7·3%), 68 medical students (6·8%), and 78 nurses and other healthcare professionals (7·8%), and 126 (12·6%) indeterminate. Of the 1000 randomly-sampled contributors, 614 were from North America (61·4%), 100 were from Europe (10·0%), 26 were from Australia or New Zealand (2·6%), 24 were from Asia (2·4%), 6 were from South America (0·6%), 5 were from the Middle East (0·5%), 4 were from Africa (0·4%), and 221 (22·1%) indeterminate. Of the 19 contributors who produced tweets flagged as blatantly false or misleading, 2 (10·5%) were staff physicians, 3 (15·8%) were organization or institutions, 2 were nurses (10·5%), 1 (5·3%) was a non-clinician researcher, 1 (5·3%) was a non-healthcare professional, and 10 (47·4%) were undetermined. Their median number of followers was 728 (IQR 189, 1145).

**Table 3:** Demographics of 1000 randomly selected contributors

| Source/Profession | Number (%) |
| --- | --- |
| Physician | 437 (43.7) |
| Not Reported | 126 (12.6) |
| Institution or Organization | 114 (11.4) |
| Resident Physician | 77 (7.7) |
| Non-healthcare professional | 73 (7.3) |
| Medical student | 68 (6.8) |
| Other Healthcare Professional | 45 (4.5) |
| Nurse | 31 (3.1) |
| Non-clinician researcher | 27 (2.7) |
| Pharmacist | 2 (0.2) |
| **Location of Contributor** |  |
| North America | 614 (61.4) |
| Not Reported | 221 (22.1) |
| Europe | 100 (10.0) |
| Australia or New Zealand | 26 (2.6) |
| Asia | 24 (2.4) |
| South America | 6 (0.6) |
| Middle East | 5 (0.5) |
| Africa | 4 (0.4) |

### Dissemination of original research

There were 275 tweets that linked to 157 unique peer reviewed original research articles. Of these, 23 were published before January 1^st^, 2020 and were excluded from analysis. The 134 remaining studies included 18 epidemiological studies (13·4%), 4 intervention studies (3·0%), 4 diagnostic studies (3·0%), 10 basic science studies (7·5%), and 98 case series or other (73·1%). The top 5 most common countries of corresponding authors were China (57, 42·5%), United States of America (29, 21·6%), Australia (11, 8·2%), United Kingdom (15, 11·2%), and Italy (8, 6·0%). The top 5 most common journals of publication were The Lancet (20, 14·9%), The Journal of the American Medical Association (18, 13·4%), The Lancet Respiratory Medicine (9, 6·7%), The New England Journal of Medicine (8, 6·0%), and The Medical Journal of Australia (8, 6·0%). The median number of days between publication and tweet was 2 (IQR 1, 5). A table with the individual studies is included in the supplementary materials.

### Dissemination of FOAM resources

There were 1122 tweets containing links to FOAM resources (websites, blogs, infographics, or attached files). The top 10 FOAM resources included in original tweets were “onepagericu.com” (47, 4·2%), “emcrit.org” (32, 2·9%), “esicm.org” (28, 2·5%), “sccm.org” (23, 2·0%), “butterflynetwork.com” (17, 1·5%), “rebelem.com” (13, 1·2%), “elso.blog” (12, 1·1%) “insightplus.mja.com.au” (12, 1·1%), “ultrasoundtraining.com.au (12, 1·1%), “intensiveblog.com (11, 1·0%), and “propofology.com” (11, 1·0%). The median number of days from publication of a FOAM resource to dissemination on Twitter was 1 (IQR 0, 3). Many of the resources that were shared were singular attachments that did not contain a link to a FOAM website, blog or resource. These included 79 documents (eg, hospital Covid-19 protocols), 83 unsourced infographics, and 27 webinars (including participant notes). The median number of days between publication of FOAM resources and tweet was shorter than for original peer reviewed research (1 vs. 2, p < 0·0001)

## DISCUSSION

Covid-19 is not the first pandemic where Twitter has played an important role in sharing information. The H1N1 pandemic and Ebola epidemic were widely discussed on Twitter, with millions of original tweets.[16,17] When compared to the H1N1 pandemic in 2009, however, the number of tweets discussing Covid-19 tweets has increased exponentially, surging in early March 2020 (Figure 2).[17] For context, at that time Europe had Covid-19 cases across the continent, with several hundred deaths in Italy alone.[18] North America also had its first significant outbreak in Washington, USA.[19] Although the FOAM community has contributors worldwide, most tweets were created by North American contributors, which may account for the temporal association between Twitter use and western outbreaks of Covid-19.

Given the immediacy and reach of social media, the FOAM community may be ideally situated to share medical resources during a pandemic. We found that more than 1 in 4 tweets contained a link to a Covid-19 resource. Compared to traditional publication peer review, the publication and editorial processes of FOAM resources varies.[14,20,21] FOAM relies on transparent and open post-publication peer review where other contributors can discuss, critique, and sometimes even contribute to resources.[8,11] One illustrations of effective post-publication peer review during the Covid-19 pandemic has been the Internet Book of Critical Care (IBCC) chapter on Covid-19.[22] From March 2^nd^ to April 16^th^, 2020 the IBCC received over 2·1 million views, with over 180 comments contributing to post publication peer review.[23] This has led to numerous revisions of the chapter to incorporate new evidence. To consolidate and share the vast amount of information being generated during a pandemic, open post-publication peer review may help balance timely dissemination of content whilst ensuring its accuracy and quality.

In addition to the speed and reach of FOAM publication, social media may be particularly effective in sharing tacit knowledge.[12,13] This stems in part from its ability to facilitate discussions and story-telling, which are key components of tacit knowledge translation.[13] During Covid-19, the geographic progression of disease from Asia, to Europe, to North America allowed for clinicians to share their experiences managing Covid-19 patients. In total, 9·8% percent of original tweets shared learned experiences and 10·8% represented individuals asking for crowd-sourced advice. Many of these early tweets pertained to the diagnosis of Covid-19 infection, ultrasound use, airway management, personal protective equipment, and mechanical ventilation. These resources may have helped clinicians and organizations to prepare for Covid-19.

With the unprecedented amount of new Covid-19 research being published, it seems increasingly difficult to find accurate and reliable content online. This has been termed an “infodemic’ by the WHO.[24] Although Twitter might contribute to this through the indiscriminate sharing of information, it can also flag important new research and give clinicians a forum to openly critique it. In our study, we found that contributors rapidly tweeted new Covid-19 research, with a median time between publication and tweet of 2 days. The majority were published in high-impact journals and included important early studies on Covid-19. The immediacy of discussion and rebuttal provided through Twitter also may be valuable, especially when many studies are first being published as pre-prints and have not undergone peer review. For example, when Gautret et al. published their initial pre-print advocating for the use of Hydroxychloroquine and Azithromycin as a treatment for Covid-19,[25] many FOAM contributors called for more rigorous studies with patient-important outcomes before widespread adoption (Table 2). Their criticisms were substantiated with recent studies showing increased adverse events and a potential association with mortality for hydroxychloroquine.[26,27]

Contributing to and participating in the FOAM community is not without risks and the unwritten rule is *caveat emptor* (buyer beware). Across social media, the potential for receiving misinformation is real and significant.[28] In this study, 19 contributors contributed blatantly false or misleading information; however, this represented only 0·2% of the total number of tweets in this analysis. Whereas blatantly misleading tweets are relatively easy to identify, a significant concern is when a reader is misled through either misrepresentation of opinion as fact, sensational anecdotes, or providing content without context. A recent study Kouzy *et al*. found that 1 in 4 tweets about Covid-19 across Twitter (no FOAM hashtags) contained misinformation.[29] We suspect the rate of subtle misinformation in the FOAM community is higher than the 0·2% found in this study; however, given that the community’s collective goal is to share legitimate knowledge, it is likely lower than the broader Twitter community.

FOAM has the potential to decrease the knowledge translation gap during Covid-19; however, resources may be of variable quality.[30] Readers are responsible for critically appraising online content; however, locating quality resources to begin with can be a challenge. The Social Media Index (SMI) provides a list of FOAM websites that are both impactful [31] and high quality,[32] analogous in some ways to a journal’s impact factor. When reading these resources, the Academic Life in Emergency Medicine (ALiEM) AIR tool or revised METRIQ tool have been validated to help determine the quality of content.[33,34]

### Limitations

Our study has several limitations. Although a broad hashtag search strategy was used, some FOAM hashtags were not included. As well, many Twitter contributors do not add hashtags to all of their Tweets, meaning that some tweets that would have been relevant were missed. Additionally, although the search was up to date as of March 21^st^, 2020, the necessary time for analysis and manuscript preparation means it does not reflect current Twitter use. As well, interrater reliability was only fair, likely related to the fact some tweets fell into several categories. This may account for the reason why a fair proportion of tweets were classified as “other Covid-19”. Finally, during initial months of Covid-19 the largest number of cases were in Asia and Europe,[1,35] yet the study was limited to English tweets, potentially selecting for a North American or Anglo-biased perspective.

## Conclusions

In the age of social media, many clinicians use Twitter to share resources and ideas with the goal of improving care for their patients. Twitter is effective in disseminating information; however, it comes with challenges in ensuring content is accurate and relevant. This study represents a first step in understanding Twitter use among the FOAM community during the Covid-19 pandemic. Further work is required to improve Twitter as a knowledge translation tool both for Covid-19 and future global crises, such that misinformation and bias is minimized, and factual knowledge dissemination maximized. Covid-19 has united clinicians around the world, and perhaps more than ever, effective strategies for sharing new ideas, accurate information, and quality research are needed.

## Data Availability

Our protocol was registered on the Open Science Framework (OSF) prior to initiation of data collection (https://osf.io/3tx96/). The original data is also published on OSF.

## COMPETING INTRESTS

None of the authors have any financial, professional, or personal competing interests to declare.

## ACKNOWLEDGEMENTS

All authors contributed in the design of the study, interpretation of data, and manuscript writing. RP, MP, SB, KW, NN also performed data analysis. RU and RP performed literature search and figure drafting. RP takes full responsibility for the integrity of the data. All authors have had access to the data and have reviewed the final manuscript.

## DATA SHARING

All data collected from this study are available publicly on the social media site Twitter.com. The specific tweets analyzed with direct links to their source material is available on OSF at https://osf.io/3tx96/. As these tweets are publicly available, no deidentification is needed. The protocol for this study was registered on the OSF prior to initiating data collection. All of these data are openly accessible by any reader without need for investigator support with no end date.

