## Supplemental tables for "Content analysis and characterization of medical tweets during the early Covid-19 pandemic"

|  | **Week 1** | **Week 2** | **Week 3** | **Week 4** | **Week 5** | **Week 6** | **Week 7** | **Week 8** | **Week 9** | **Total Tweets** | **P value*** |
| --- | --- | --- | --- | --- | --- | --- | --- | --- | --- | --- | --- |
| **Original Research** | 1  (1.6%) | 2 (2.3%) | 5 (8.1%) | 2 (2.6%) | 7 (9.1%) | 10 (5.1%) | 28 (4.1%) | 61 (2.6%) | 159 (3.2%) | 275 | 0.47 |
| **Editorials** | 2 (3.3%) | 6 (6.9%) | 6 (9.7%) | 7 (9.1%) | 8 (10.4%) | 12 (6.1%) | 27 (4.0%) | 74 (3.2%) | 162 (3.2%) | 304 | 0.25 |
| **FOAM Content** | 7 (11.5%) | 14 (16.1%) | 9 (14.5%) | 17 (22.1%) | 10 (13.0%) | 14 (7.1%) | 104 (15.4%) | 252 (10.9%) | 695 (13.9%) | 1122 | 0.6 |
| **Public Health** | 1 (1.6%) | 8 (9.2%) | 1 (1.6%) | 3 (3.9%) | 3 (3.9%) | 13 (6.6%) | 26 (3.9%) | 57 (2.5%) | 133 (2.7%) | 245 | 0.83 |
| **Podcast or Video** | 2 (3.3%) | 3 (3.4%) | 4 (6.5%) | 4 (5.2%) | 5 (6.5%) | 3 (1.5%) | 35 (5.2%) | 88 (3.8%) | 266 (5.3%) | 410 | 0.6 |
| **Learned Experience** | 4 (6.6%) | 8 (9.2%) | 5 (8.1%) | 4 (5.2%) | 4 (5.2%) | 20 (10.2%) | 50 (7.4%) | 247 (10.7%) | 531 (10.6%) | 873 | 0.21 |
| **Refuting False Informatino** | 0 (0.0%) | 4 (4.6%) | 4 (6.5%) | 2 (2.6%) | 2 (2.6%) | 1 (0.5%) | 10 (1.5%) | 34 (1.5%) | 26 (0.5%) | 83 | 0.29 |
| **Policy Discussion** | 7 (11.5%) | 13 (14.9%) | 5 (8.1%) | 10 (13.0%) | 6 (7.8%) | 26 (13.3%) | 79 (11.7%) | 275 (11.9%) | 690 (13.8%) | 1111 | 0.6 |
| **Emotional Impact** | 7 (11.5%) | 9 (10.3%) | 7 (11.3%) | 8 (10.4%) | 9 (11.7%) | 23 (11.7%) | 67 (9.9%) | 376 (16.2%) | 684 (13.7%) | 1190 | 0.18 |
| **Blatantly False** | 0 (0.0%) | 0 (0.0%) | 0 (0.0%) | 0 (0.0%) | 2 (2.6%) | 1 (0.5%) | 4 (0.6%) | 4 (0.2%) | 8 (0.2%) | 19 | 0.23 |
| **Advice** | 6 (9.8%) | 2 (2.3%) | 1 (1.6%) | 2 (2.6%) | 3 (3.9%) | 31 (15.8%) | 74 (11.0%) | 321 (13.9%) | 488 (9.8%) | 928 | 0.23 |
| **Other Covid** | 20 (32.8%) | 13 (14.9%) | 14 (22.6%) | 14 (18.2%) | 16 (20.8%) | 32 (16.3%) | 136 (20.1%) | 404 (17.5%) | 908 (18.2%) | 1557 | 0.18 |
| **Non-Covid** | 4 (6.6%) | 5 (5.7%) | 1 (1.6%) | 4 (5.2%) | 2 (2.6%) | 10 (5.1%) | 35 (5.2%) | 122 (5.3%) | 241 (4.8%) | 424 | 0.47 |
| **Total Tweets Per Week** | 61 | 87 | 62 | 77 | 77 | 196 | 675 | 2315 | 4991 | 8541 |  |
| *Man Kendall Test for Monotonic Trends | | | | | | | | | | | |

**Supplemental Table 1:** Number of original tweets by content category per week

**Supplementary Table 2:** Research articles with Pubmed ID linked in original tweets

| **PMID** | **Study Title** | **Journal of Publication** | **Article Type*** |
| --- | --- | --- | --- |
| 25599463 | Fluid Management With a Simplified Conservative Protocol for the Acute Respiratory Distress Syndrome | Critical Care Medicine | 1 |
| 25903751 | A cluster randomised trial of cloth masks compared with medical masks in healthcare workers | BMJ Open | 1 |
| 29071828 | Intactness of Medical Nonsterile Gloves on Use of Alcohol Disinfectants | Ann Lab Med | 1 |
| 30336170 | Comparison of high-flow nasal cannula versus oxygen face mask for environmental bacterial contamination in critically ill pneumonia patients: a randomized controlled crossover trial | Journal of Hospital Infection | 1 |
| 31479137 | N95 Respirators vs Medical Masks for Preventing Influenza Among Health Care Personnel | JAMA | 1 |
| 32178954 | Anesthetic Management of Patients with COVID 19 Infections during Emergency Procedures | Journal of Cardiothoracic & Vascular Anesthesia | 1 |
| 32187464 | A Trial of Lopinavir–Ritonavir in Adults Hospitalized with Severe Covid-19 | NEJM | 1 |
| 32205204 | Hydroxychloroquine and azithromycin as a treatment of COVID-19: results of an open-label non-randomized clinical trial | International Journal of  Antimicrobial Agents | 1 |
| 32049601 | Chest CT for Typical 2019-nCoV Pneumonia: Relationship to Negative RT-PCR Testing | Radiology | 2 |
| 32166346 | Findings of lung ultrasonography of novel corona virus pneumonia during the 2019–2020 epidemic | Intensive Care Medicine | 2 |
| 16115318 | Chloroquine is a potent inhibitor of SARS coronavirus infection and spread | Virology Journal | 3 |
| 17302372 | Indomethacin has a potent antiviral activity against SARS coronavirus. | Antiviral therapy | 3 |
| 17522231 | Clathrin-Dependent Entry of Severe Acute Respiratory Syndrome Coronavirus into Target Cells Expressing ACE2 with the Cytoplasmic Tail Deleted | Journal of Virology | 3 |
| 20430477 | Inactivation of influenza A virus H1N1 by disinfection process. | Am J Infect Control | 3 |
| 22436202 | Cough aerosol in healthy participants: fundamental knowledge to optimize droplet-spread infectious respiratory disease management | BMC Pulmonary Medicine | 3 |
| 25224111 | Aerosol Dispersion During Various Respiratory Therapies: A Risk Assessment Model of Nosocomial Infection to Health Care Workers | Hong Kong Med J | 3 |
| 28193191 | Effective inhibition of MERS-CoV infection by resveratrol | BMC Infectious Diseases | 3 |
| 29511076 | Coronavirus Susceptibility to the Antiviral Remdesivir (GS-5734) Is Mediated by the Viral Polymerase and the Proofreading Exoribonuclease | American Society for Microbiology | 3 |
| 29678452 | Ultraviolet Germicidal Irradiation of Influenza-Contaminated N95 Filtering Facepiece Respirators | Am J Infect Control | 3 |
| 31753725 | Pro-inflammatory monocyte profile in patients with major depressive disorder and suicide behaviour and how ketamine induces anti-inflammatory M2 macrophages by NMDAR and mTOR | EBio Medicine | 3 |
| 32020029 | Remdesivir and chloroquine effectively inhibit the recently emerged novel coronavirus (2019-nCoV) in vitro | Cell Research | 3 |
| 32075877 | Cryo-EM structure of the 2019-nCoV spike in the prefusion conformation | Science | 3 |
| 32150618 | In Vitro Antiviral Activity and Projection of Optimized Dosing Design of Hydroxychloroquine for the Treatment of Severe Acute Respiratory Syndrome Coronavirus 2 (SARS-CoV-2) | Clinical Infectious Diseases | 3 |
| 32182409 | Aerosol and surface stability of HCoV-19 (SARS-CoV-2) compared to SARS-CoV-1 | NEJM | 3 |
| 32192578 | COVID-19: consider cytokine storm syndromes and immunosuppression | The Lancet | 3 |
| 32194981 | Hydroxychloroquine, a less toxic derivative of chloroquine, is effective in inhibiting SARS-CoV-2 infection in vitro | Cell Discovery | 3 |
| 32199469 | Prolonged presence of SARS-CoV-2 viral RNA in faecal samples | Lancet Gastroenterology & Hepatology | 3 |
| 32237278 | Isolation and rapid sharing of the 2019 novel coronavirus (SARS-CoV-2) from the first patient diagnosed with COVID-19 in Australia | Medical Journal of Australia | 3 |
| 25637115 | Face touching: a frequent habit that has implications for hand hygiene. | Am J Infect Control | 4 |
| 31585142 | Mobile phones as fomites for potential pathogens in hospitals: microbiome analysis reveals hidden contaminants | Journal of Hospital Infection | 4 |
| 31607599 | Influenza vaccination and respiratory virus interference among Department of Defense personnel during the 2017-2018 influenza season. | Vaccine | 4 |
| 32064853 | The epidemiological characteristics of an outbreak of 2019 novel coronavirus diseases (COVID-19) in China | CMAPH | 4 |
| 32105632 | Clinical course and outcomes of critically ill patients with SARS-CoV-2 pneumonia in Wuhan, China: a single-centered, retrospective, observational study | Lancet Respiratory Medicine | 4 |
| 32119825 | Feasibility of controlling COVID-19 outbreaks by isolation of cases and contacts | Lancet Global Health | 4 |
| 32167524 | Risk Factors Associated With Acute Respiratory Distress Syndrome and Death in Patients With Coronavirus Disease 2019 Pneumonia in Wuhan, China | JAMA Internal Medicine | 4 |
| 32171076 | Clinical course and risk factors for mortality of adult inpatients with COVID-19 in Wuhan, China: a retrospective cohort study | Lancet | 4 |
| 32171390 | Real estimates of mortality following COVID-19 infection | Lancet | 4 |
| 32179660 | Epidemiology of COVID-19 Among Children in China | Pediatrics | 4 |
| 32179701 | Substantial undocumented infection facilitates the rapid dissemination of novel coronavirus (SARS-CoV2) | Science | 4 |
| 32179890 | Risk Factors of Healthcare Workers with Corona Virus Disease 2019: A Retrospective Cohort Study in a Designated Hospital of Wuhan in China | Clinical Infectious Diseases | 4 |
| 15963157 | The Impact of Severe Acute Respiratory Syndrome on Medical House Staff | J Gen Intern Med | 5 |
| 16400088 | Concept of Operations for Triage of Mechanical Ventilation in an Epidemic | Academic Emergency Medicine | 5 |
| 16885402 | A Single Ventilator for Multiple SimulatedPatients to Meet Disaster Surge | Academic Emergency Medicine | 5 |
| 19773323 | Physical 1s to interrupt or reduce the spread of respiratory viruses: systematic review | BMJ | 5 |
| 23538558 | Search for inhibitors of endocytosis | Cellular Logistics | 5 |
| 28556555 | Examination of Hydroxychloroquine Use and Hemolytic Anemia in G6PDH-Deficient Patients | Arthritis Care Res | 5 |
| 28586113 | Lung Consolidation Locations for Optimal Lung Ultrasound Scanning in Diagnosing Pediatric Pneumonia | J Ultrasound Med | 5 |
| 30368986 | Lung‐protective Ventilation for Acute Respiratory Distress Syndrome | Academic Emergency Medicine | 5 |
| 31766051 | Standard and Precordial Leads Obtained With an Apple Watch | Annals of Internal Medicine | 5 |
| 31871560 | Involving Physicians-in-Training in the Care of Patients During Epidemics | Journal of Graduate Medical Education | 5 |
| 31967327 | Emerging coronaviruses: Genome structure, replication, and pathogenesis | Journal of Medical Virology | 5 |
| 31986264 | Clinical features of patients infected with 2019 novel coronavirus in Wuhan, China | Lancet | 5 |
| 32004427 | First Case of 2019 Novel Coronavirus in the United States | NEJM | 5 |
| 32022836 | 2019 Novel Coronavirus—Important Information for Clinicians | JAMA | 5 |
| 32031570 | Clinical Characteristics of 138 Hospitalized Patients With 2019 Novel Coronavirus–Infected Pneumonia in Wuhan, China | JAMA | 5 |
| 32034323 | Mechanisms of action of hydroxychloroquine and chloroquine: implications for rheumatology | Nature Reviews | 5 |
| 32035030 | The mental health of medical workers in Wuhan, China dealing with the 2019 novel coronavirus | Lancet | 5 |
| 32052373 | Practical recommendations for critical care and anesthesiology teams caring for novel coronavirus (2019-nCoV) patients | Canadian Journal of Anesthesia | 5 |
| 32061335 | Therapeutic and triage strategies for 2019 novel coronavirus disease in fever clinics | Lancet Respiratory Medicine | 5 |
| 32066488 | Validation of neuromuscular blocking agent use in acute respiratory distress syndrome: a meta-analysis of randomized trials | Critical Care | 5 |
| 32066541 | Cancer patients in SARS-CoV-2 infection: a nationwide analysis in China | Lancet Oncology | 5 |
| 32074258 | Preparing for the Most Critically Ill Patients With COVID-19 The Potential Role of Extracorporeal Membrane Oxygenation | JAMA | 5 |
| 32077115 | Clinical characteristics of 140 patients infected with SARS‐CoV‐2 in Wuhan, China | Allergy | 5 |
| 32085846 | Pathological findings of COVID-19 associated with acute respiratory distress syndrome | Lancet Respiratory Medicine | 5 |
| 32085849 | The first Vietnamese case of COVID-19 acquired from China | Lancet Infectious Diseases | 5 |
| 32087116 | Asymptomatic cases in a family cluster with SARS-CoV-2 infection | Lancet Infectious Diseases | 5 |
| 32091533 | Characteristics of and Important Lessons From the Coronavirus Disease 2019 (COVID-19) Outbreak in China | JAMA | 5 |
| 32100024 | De-isolating Coronavirus Disease 2019 Suspected Cases: A Continuing Challenge | Clinical Infectious Diseases | 5 |
| 32101510 | Correlation of Chest CT and RT-PCR Testing in Coronavirus Disease 2019 (COVID-19) in China: A Report of 1014 Cases | Radiology | 5 |
| 32105633 | Staff safety during emergency airway management for COVID-19 in Hong Kong | Lancet Respiratory Medicine | 5 |
| 32109013 | Clinical Characteristics of Coronavirus Disease 2019 in China | NEJM | 5 |
| 32112714 | The psychological impact of quarantine and how to reduce it: rapid review of the evidence | Lancet | 5 |
| 32115364 | Epidemiological and clinical characteristics of heart transplant recipients during the 2019 coronavirus outbreak in Wuhan china; a descriptive survey report | Journal of Heart and Lung Transplantation | 5 |
| 32125362 | Epidemiologic Features and Clinical Course of Patients Infected With SARS-CoV-2 in Singapore. | JAMA | 5 |
| 32125452 | Clinical predictors of mortality due to COVID-19 based on an analysis of data of 150 patients from Wuhan, China | Intensive Care Medicine | 5 |
| 32125455 | Angiotensin-converting enzyme 2 (ACE2) as a SARS-CoV-2 receptor: molecular mechanisms and potential therapeutic target | Intensive Care Medicine | 5 |
| 32129805 | Air, Surface Environmental, and Personal Protective Equipment Contamination by Severe Acute Respiratory Syndrome Coronavirus 2 (SARS-CoV-2) From a Symptomatic Patient | JAMA | 5 |
| 32133578 | Review of the Clinical Characteristics of Coronavirus Disease 2019 (COVID-19) | J Gen Intern Med | 5 |
| 32134381 | Lack of Vertical Transmission of Severe Acute Respiratory Syndrome Coronavirus 2, China | Emerging Infectious Diseases | 5 |
| 32144127 | Coronavirus disease 2019 (covid-19): a guide for UK GPs | BMJ | 5 |
| 32145829 | Respiratory support for patients with COVID-19 infection | Lancet | 5 |
| 32146721 | An outbreak of COVID‐19 caused by a new coronavirus: what we know so far | Medical Journal of Australia | 5 |
| 32150360 | Features, Evaluation and Treatment Coronavirus (COVID-19) | | 5 |
| 32150748 | The Incubation Period of Coronavirus Disease 2019 (COVID-19) From Publicly Reported Confirmed Cases: Estimation and Application | Annals of Internal Medicine | 5 |
| 32151335 | Clinical characteristics and intrauterine vertical transmission potential of COVID-19 infection in nine pregnant women: a retrospective review of medical records | Lancet | 5 |
| 32159735 | Care for Critically Ill Patients With COVID-19 | JAMA | 5 |
| 32160316 | Co‐infection of SARS‐CoV‐2 and HIV in a patient in Wuhancity, China | Journal of Medical Virology | 5 |
| 32163697 | Detection of Covid-19 in Children in Early January 2020 in Wuhan, China | NEJM | 5 |
| 32166318 | Protecting Health Care Workers During the COVID-19 Coronavirus Outbreak -Lessons From Taiwan's SARS Response | Clinical Infectious Diseases | 5 |
| 32166346 | Findings of lung ultrasonography of novel corona virus pneumonia during the 2019–2020 epidemic | Intensive Care Medicine | 5 |
| 32167525 | From Containment to Mitigation of COVID-19 in the US | JAMA | 5 |
| 32167538 | Critical Care Utilization for the COVID-19 Outbreak in Lombardy, Italy: Early Experience and Forecast During an Emergency Response | JAMA | 5 |
| 32167853 | Can Lung US Help Critical Care Clinicians in the Early Diagnosis of Novel Coronavirus (COVID-19) Pneumonia? | Radiology | 5 |
| 32171062 | Are patients with hypertension and diabetes mellitus at increased risk for COVID-19 infection? | Lancet Respiratory Medicine | 5 |
| 32172175 | Clinical considerations for patients with diabetes in times of COVID-19 epidemic | Diabetes & Metabolic Syndrome: Clinical Research & Reviews | 5 |
| 32173110 | A systematic review on the efficacy and safety of chloroquine for the treatment of COVID-19 | Journal of Critical Care | 5 |
| 32176257 | COVID-19 and the Risk to Health Care Workers: A Case Report | Annals of Internal Medicine | 5 |
| 32180175 | Safety and efficacy of different anesthetic regimens for parturients with COVID-19 undergoing Cesarean delivery: a case series of 17 patients | Canadian Journal of Anesthesia | 5 |
| 32180426 | An Analysis of 38 Pregnant Women with COVID-19, Their Newborn Infants, and Maternal3 Fetal Transmission of SARS-CoV-2: Maternal Coronavirus Infections and Pregnancy Outcomes | Archives of Pathology & Laboratory Medicine | 5 |
| 32181795 | Coronavirus Disease 2019 (COVID-19) in Italy | JAMA | 5 |
| 32182347 | Perinatal Transmission of COVID-19 Associated SARS-CoV-2: Should We Worry? | Clinical Infectious Diseases | 5 |
| 32187458 | SARS-CoV-2 Infection in Children | NEJM | 5 |
| 32191259 | Characteristics and Outcomes of 21 Critically Ill Patients With COVID-19 in Washington State | JAMA | 5 |
| 32192578 | COVID-19: consider cytokine storm syndromes and immunosuppression | Lancet | 5 |
| 32199075 | Health security capacities in the context of COVID-19 outbreak: an analysis of International Health Regulations annual report data from 182 countries | Lancet | 5 |
| 32199938 | Catheterization Laboratory Considerations During the Coronavirus (COVID-19) Pandemic: From ACC’s 1al Council and SCAI | Journal of the American College of Cardiology | 5 |
| 32203711 | Planning and provision of ECMO services for severe ARDS during the COVID-19 pandemic and other outbreaks of emerging infectious diseases | Lancet Respiratory Medicine | 5 |
| 32204907 | The Novel Coronavirus 2019 epidemic and kidneys | Kidney International | 5 |
| 32224769 | Surviving Sepsis Campaign: Guidelines on the Management of Critically Ill Adults with Coronavirus Disease 2019 (COVID-19) | Intensive Care Medicine | 5 |
| 32237278 | Isolation and rapid sharing of the 2019 novel coronavirus (SARS-CoV-2) from the first patient diagnosed with COVID-19 in Australia | Medical Journal of Australia | 5 |
| 32266965 | COVID-19 precautions – easier said than done when patients are homeless | Medical Journal of Australia | 5 |
| 32275288 | Neurological Manifestations of Hospitalized Patients with COVID-19 in Wuhan, China: a retrospective case series study | N/A (medRxiv) | 5 |
| *Category:  1 = Intervention  2 = diagnostic  3 = basic science  4 = epidemiological  5= case studies, series, or other | | | |
